# The Impact of COVID-19 Vaccination on California’s Return to *Normalcy*

**DOI:** 10.1101/2021.06.01.21258187

**Authors:** Maria L. Daza–Torres, Yury E. García, Alec J. Schmidt, Brad H Pollock, James Sharpnack, Miriam Nuño

**Affiliations:** Department of Public Health Sciences, University of California Davis, CA, USA; Department of Statistics, University of California Davis, CA, USA

## Abstract

SARS-CoV-2 has infected nearly 3.7 million and killed 61,722 Californians, as of May 22, 2021. Non-pharmaceutical interventions have been instrumental in mitigating the spread of the coronavirus. However, as we ease restrictions, widespread implementation of COVID- 19 vaccines is essential to prevent its resurgence. In this work, we addressed the adequacy and deficiency of vaccine uptake within California and the possibility and severity of resurgence of COVID-19 as restrictions are lifted given the current vaccination rates. We implemented a real-time Bayesian data assimilation approach to provide projections of incident cases and deaths in California following the reopening of its economy on June 15, 2021. We implemented scenarios that vary vaccine uptake prior to reopening, and transmission rates and effective population sizes following the reopening. For comparison purposes, we adopted a baseline scenario using the current vaccination rates, which projects a total 11,429 cases and 429 deaths in a 15-day period after reopening. We used posterior estimates based on CA historical data to provide realistic model parameters after reopening. When the transmission rate is increased after reopening, we projected an increase in cases by 21.8% and deaths by 4.4% above the baseline after reopening. When the effective population is increased after reopening, we observed an increase in cases by 51.8% and deaths by 12.3% above baseline. A 30% reduction in vaccine uptake alone has the potential to increase cases and deaths by 35% and 21.6%, respectively. Conversely, increasing vaccine uptake by 30% could decrease cases and deaths by 26.1% and 17.9%, respectively. As California unfolds its plan to reopen its economy on June 15, 2021, it is critical that social distancing and public behavior changes continue to be promoted, particularly in communities with low vaccine uptake. The Centers of Disease Control’s (CDC) recommendation to ease mask-wearing for fully vaccinated individuals despite major inequities in vaccine uptake in counties across the state highlights some of the logistical challenges that society faces as we enthusiastically phase out of this pandemic.

## Introduction

The coronavirus pandemic has highlighted how inadequate and unprepared the public health and healthcare system was for such an event, with disproportionate consequences for traditionally underserved populations. As of May 19, 2021, the US had administered at least one dose of available vaccine to nearly 48% of the adult population, with 37.8% fully vaccinated. California has used 79% of its vaccine supplies to fully vaccinate 47.8% of the adult population, ranking 27th, proportionally, out of all states.

Management of the pandemic at the county level has thus far been lead by California’s *Blueprint for a Safer Economy* [3], which uses local case and test positivity rates, adjusted for equity measures in the most vulnerable census tracts, to determine a color-based qualitative threat level for the entire county. Business operation, non-pharmaceutical intervention (NPI) mandates, and general advice on safety and maximum gathering sizes are tied to these threat levels. In the recent *Beyond the Blueprint for a Safer Economy*, The California Department of Public Health (CDPH) issued plans and guidance for a full easing of all business restrictions by June 15th, 2021, contingent on case, vaccination, and equity measurements staying on track. CDPH stressed the importance of maintaining infrastructure and resources for continuing vaccination programs; monitoring for new cases and strains with active testing, especially in the most vulnerable areas of the state; robust contact tracing and outbreak investigations; statewide plans to scale up resources in the case of another large outbreak; and monitoring hospital usage, appropriate availability of personal protective equipment, and surge capacity [2].

It is unlikely that any given state in the US will be able to eliminate SARS-CoV-2 completely. Short of elimination, the next best hope is that forcing the virus into endemicity will result in a much less severe disease in years to come [20], which is the foundation for California’s current approach. For infectious diseases like COVID-19, the risk and size of an outbreak— and the threshold for forced endemic transmission—is determined by the transmissibility of the virus and the effective size of the population who can acquire it. Transmissibility is a function of biochemical, biological, and social factors and can thus change over time in unpredictable ways. Multiple new strains of the SARS-CoV-2 virus have already appeared over the course of this pandemic, the interactions of which with natural and pharmaceutical immunity is only partially explored [19]. Though acquiring an infection after a complete vaccination regimen is rare [18, 23], the continued appearance of new strains, the risk of which increases with circulation around the world, could reduce vaccine effectiveness [5, 6]. Additionally, messaging around the lifted restrictions and pandemic fatigue may lead people to change their behaviors in ways that could change transmission rates, such as ignoring mask use, social distancing, and limitations on indoor gatherings of unvaccinated individuals [1, 8].

The effective susceptible population includes all who have not recovered from the virus, those whose immunity has waned and thus may acquire the virus again, and those rarer individuals who have a breakthrough event from a vaccine failure. Each of these categories are difficult to estimate, especially in the context of how they can influence the severity of outbreaks in the future. Waning immunity, which is still poorly understood in both natural and pharmaceutical cases, could be adding more people to the susceptible pool than previously expected. Additionally, as more people feel comfortable socializing in public, there may be a larger pool of people who may not have been exposed had limitations on indoor gatherings been maintained.

Traditional SEIR models with fixed model parameters tend to be too simplistic or divergent from real-world data to act as good assessments of future trends to form the basis of an exit strategy. As an alternative approach, we implemented a Bayesian sequential data assimilation model that allows for non-stationarity by sequentially updating model parameters, such as transmission rates and the effective population. In this way, our model can implicitly account for the unobserved changes in social distancing behavior, mobility, masking behavior, etc. We used posterior coverage intervals to investigate the uncertainty of and establish reasonable ranges for these model parameters to create realistic scenarios for widescale re-opening and the lifting of restrictions. We summarize the potential impact of changes in transmissibility, effective susceptible population, and vaccination rates by observing how they can affect incident cases and deaths in the weeks after the easing of restrictions in California.

## Materials and methods

### Data Sources

Daily COVID-19 confirmed cases, deaths, and vaccination for California were obtained from *California’s open data portal* [4,16]. All data are publicly available and fully anonymized, and therefore did not require ethical approval of an institutional review board nor written informed consent. All analyses were conducted with data updated to May 18, 2021. For inference, we calculated the weekly moving average for cases and deaths.

### Mathematical Model

A dynamic SARS-CoV-2 transmission model with vaccination was implemented (Fig 1).The total population was divided into eight classes: susceptible, exposed, reported infectious, unreported contagious, vaccinated, recovered, and deceased. Individuals in the susceptible (S) group become infected and move to the exposed group with the incubation of the virus. Exposed (E) individuals subsequently move to the reported infectious group (O) or the group of unreported contagious (U). Individuals move to compartment *V*_1_ after receiving the first vaccine dose and move from *V*_1_ to *V*_2_ when fully vaccinated. Lastly, individuals are removed through recovery (R) or COVID-19 induced death (D). Once an individual is recovered, immunity is assumed for the entire period of the simulation.

**Figure 1.**
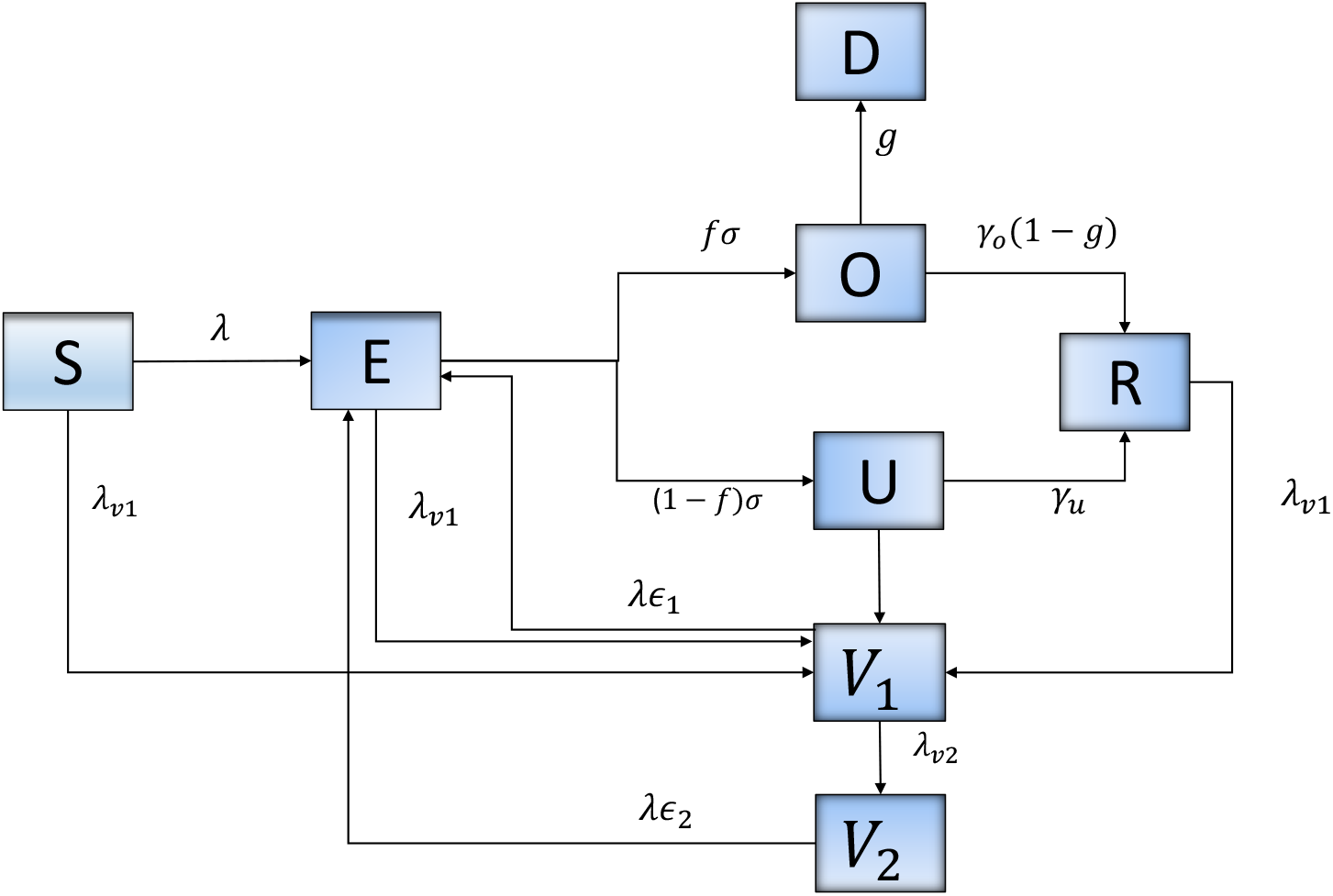
Flow diagram of SARS-CoV-2 transmission dynamics with vaccination.

### Model Structure

A dynamic model of SARS-CoV-2 transmission was developed based on eight epidemiological classes as illustrated in Fig 1. We model the process of vaccination by considering that: (1) individuals in all epidemiological classes, with the exception of those diagnosed as positive are vaccinated, (2) vaccine is partially effective, suggesting that some individuals vaccinated can be infected, (3) vaccine efficacy is different after first and second dose administration, and (4) vaccination rate is estimated using daily data of first and second doses administered in California.

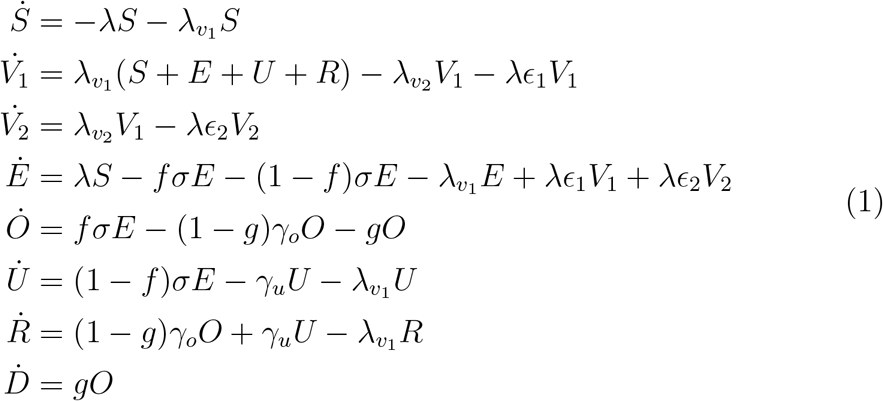

We assume that only a small proportion of individuals diagnosed as positive contribute to new infections because they do not follow recommendations to isolate. We assume that asymptomatic individuals can also be infectious, so 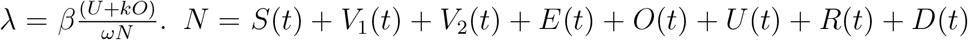 corresponds to the total population of California. Parameter description and values are summarized in Table [1].

### Parameter estimation

The Bayesian sequential forecasting method in Daza-Torres, et al. [13] is used to conduct parameter inference. The estimation is implemented by decoupling the model into two parts. First, we consider the transitions for vaccination [equation 3] and second, the remaining dynamics [equation 2].

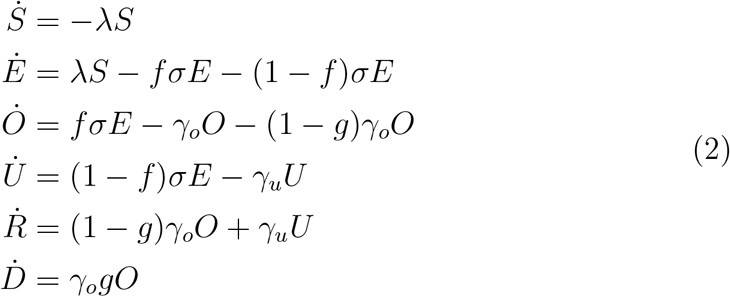

Data on weekly moving average of confirmed cases and deaths are used to estimate the contact rate (*β*), the proportion of the effective population (*ω*), the fraction of individuals infected that are deceased (*g*), and the initial conditions for all compartments, except for the susceptible ones which is set as *S*(*t*_0_) = *ω* · *N* − (*E*(*t*_0_) + *O*(*t*_0_) + *U* (*t*_0_) + *R*(*t*_0_)) + *V*_1_(*t*_0_) + *V*_2_(*t*_0_).

Let *W* be the population of non-vaccinated individuals at time *t*. For *t*_0_ = 0, *W* (*t*_0_) = *N*. We assume a vaccination rate, for first and second doses to be constant and proportional to the actual population. Therefore,

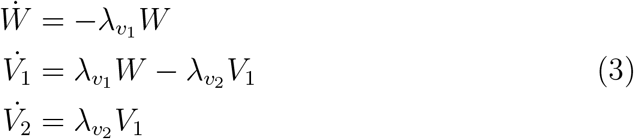

Data on individuals with at least one dose and fully vaccinated is required to find the value of 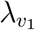 and 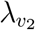.

### Scenarios

We propose a set of scenarios to analyze the impact of reopening on June 15th, considering changes in vaccination rate, virus transmission rate, and effective population. Variability in the rate of infection can inherently reflect the use of masks and new strains of the circulating virus. Changes in the effective population implicitly capture the number of people who are available to be infected due to restrictions or openings.

For the baseline scenario, we fitted the model to updated data until May 18, 2021, and we projected total positive cases and deaths from May 19 through July 15, 2021. Starting on June 15th (reopening day), simulations were evaluated with two different values for both transmission rate (*β*) and effective susceptible population (*ω*). Other parameters were the same as those estimated in the baseline scenario. Replications were evaluated for situations where the vaccination rate was reduced or increased by 30%. Table [2] summarizes the scenario assumptions.

## Results

Daily confirmed infections and deaths were projected through July 15, 2021. Fig 2 depicts the predicted daily cases and deaths for California after reopening day. The black line corresponds to the baseline scenario, the cyan line corresponds to the projection assuming a 30% reduction in the current vaccination rate, and the magenta line corresponds to the projection assuming a 30 % increment in the current vaccination rate. After June 15, the red, blue, and black lines correspond to the projection with the different values of *β* (figures (a) and (b)) and *ω* (figures (c) and (d)). Assessing the impact of different rates of transmission (*β*) allows for flexibility in changes in transmission resulting from reduction in social distancing measures, mask-wearing, and uncertain circulation of new strains after reopening. We further considered scenarios that allow for variability in the effective population size (*ω*), as it is likely that reopening will result in increased social interactions with unvaccinated, susceptible individuals. Parameter assumptions for the rate of transmission and the effective population were estimated from California’s own pandemic trajectory (Supplemental Fig S6).

**Figure 2.**
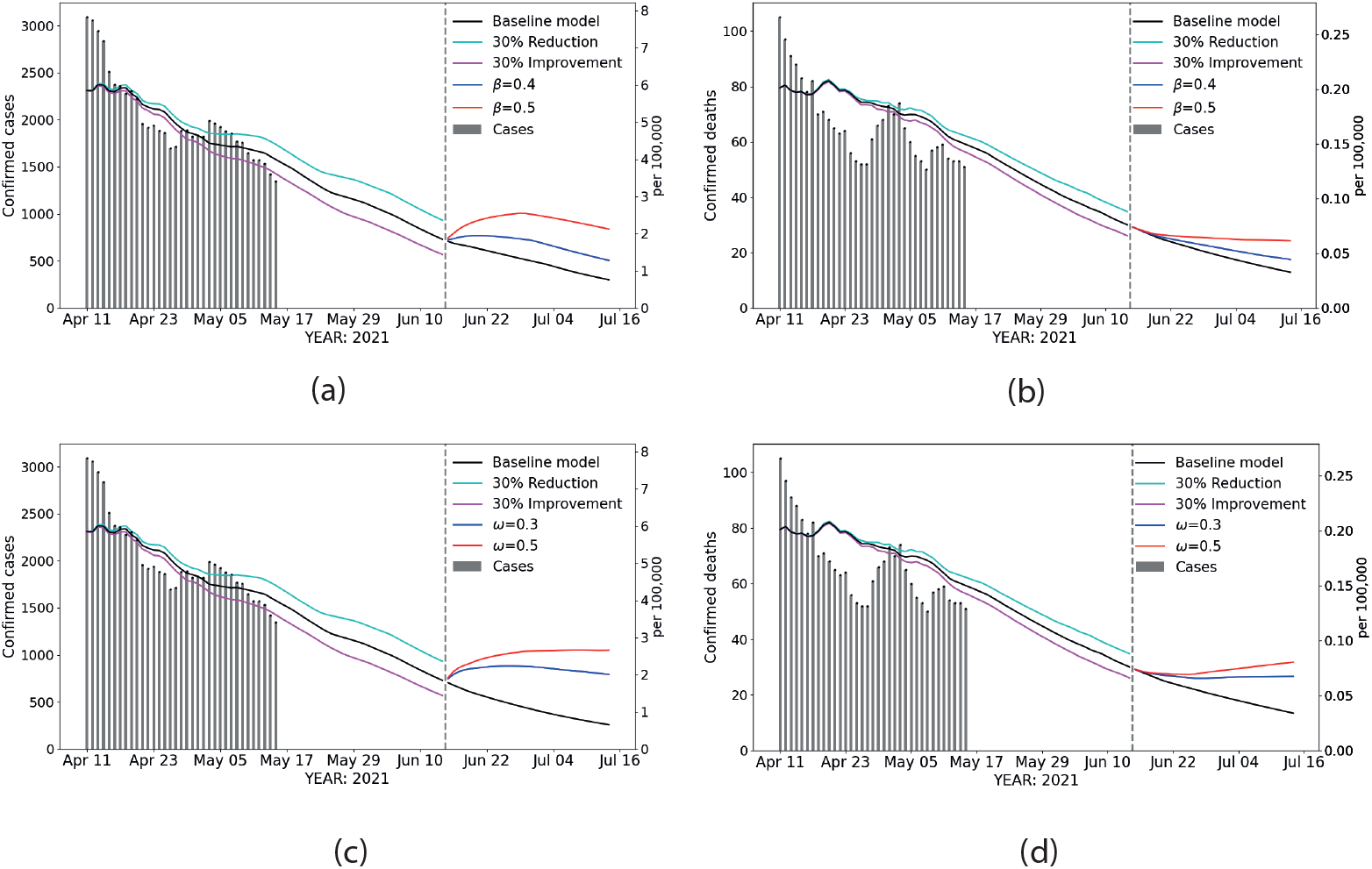
Scenarios. Estimated statewide confirmed cases and confirmed deaths during live data collection, extrapolated between May 18th and June 15th for different vaccination rates, and predicted beyond June 15th with different effective susceptible populations and transmission rates. (a) and (b): effects of varying transmission rate on confirmed cases and confirmed deaths, respectively. (c) and (d): effects of varying effective susceptible populations on confirmed cases and confirmed deaths, respectively. Dashed vertical line: June 15th. Gray vertical bars: daily reported data. Black line: baseline scenario, before and after opening. Cyan line: projection from May 18th assuming a 30% reduction in the current vaccination rate. Magenta line: projection from May 18th assuming a 30% increase in the current vaccination rate. The values used for *β* = 0.4, 0.5, and *ω* = 0.3, 0.5 were selected according to historical data in CA (Fig. S6).

### Vaccine and transmission rate effects on projections

After June 15 and assuming current vaccine uptake (*Baseline model*), confirmed cases in California approached 1 case per 100,000 population and 0.03 deaths per 100,000 population (Figure 2(a) (b)). Increasing transmission rate to 0.5, a value that reflects the highest peak experienced in California, has the potential to double cases and deaths from baseline estimates.

Table 3 provides the estimates of confirmed cases and deaths assuming changes in vaccination rates, transmission rates, and the effective population size. Assuming that the current vaccine uptake remains consistent with the current trend but we assume a transmission rate similar to the one observed during California’s highest peak, we project a 48.5% and 9.6% increase from baseline in cases and deaths, respectively, 15 days after reopening. Simultaneously increasing the rate of transmission to 0.4 and reducing vaccine rate by 30% increases the percentage change in cases by 65.7% compared to baseline. Increasing vaccination rate by 30% from the current trajectory has the potential to reduce cases (10.1%) and deaths (14.6%) even under a rate of transmission that is higher than baseline (*β*=0.31).

**Table 1:** Parameter definition and estimates.

| Parameter | Description |  |  |
| --- | --- | --- | --- |
| $\beta$ | Transmission rate | Estimated | |
| $\omega$ | Effective population | Estimated | |
| $\lambda_{v1}$ | Vaccination rate, first dose | Estimated | |
| $\lambda_{v2}$ | Vaccination rate, second dose | Estimated | |
| $g$ | Mortality rate | Estimated | |
| Parameter | Description | Value | Ref. |
| $\kappa$ | Proportion of observed people that contribute to new infections | 0.2 | |
| $f$ | Proportion of observed individuals | 0.6 | |
| $N$ | California population size | 39512223 | [7] |
| $1/\gamma_o$ | Average time to recovery, diagnosed | 1/14 | [10, 24] |
| $1/\gamma_u$ | Average time to recovery, undiagnosed | 1/7 | [22] |
| $\epsilon_1$ | $(1 - \epsilon_1)$ Vaccine efficacy after first dose | 0.40 | [12] |
| $\epsilon_2$ | $(1 - \epsilon_2)$ Vaccine efficacy after second dose | 0.05 | [12] |
| $\sigma$ | Median number of days from symptom onset | 1/5 | [14] |

**Table 2:** Summary of scenarios.

| Description | Vaccination rate assumptions |  |  |
| --- | --- | --- | --- |
|  | Current rate maintained | Current rate reduced by 30% | Current rate increased by 30% |
| Assumption: the viral transmission rate changes after June 15th. | $\beta_1 = 0.4$<br>$\beta_2 = 0.5$ | $\beta_1 = 0.4$<br>$\beta_2 = 0.5$ | $\beta_1 = 0.4$<br>$\beta_2 = 0.5$ |
| Assumption: the effective proportion of susceptible people changes after June 15th. | $\omega_1 = 0.3$<br>$\omega_2 = 0.5$ | $\omega_1 = 0.3$<br>$\omega_1 = 0.5$ | $\omega_1 = 0.3$<br>$\omega_1 = 0.5$ |

Changes in vaccination rates are paired with changes in the effective susceptible population ( $\omega$ ) or the transmission rate ( $\beta$ ). Model for the baseline scenario is fitted using updated data through May 18, 2021, generating parameters as in Table [1]

**Table 3:**
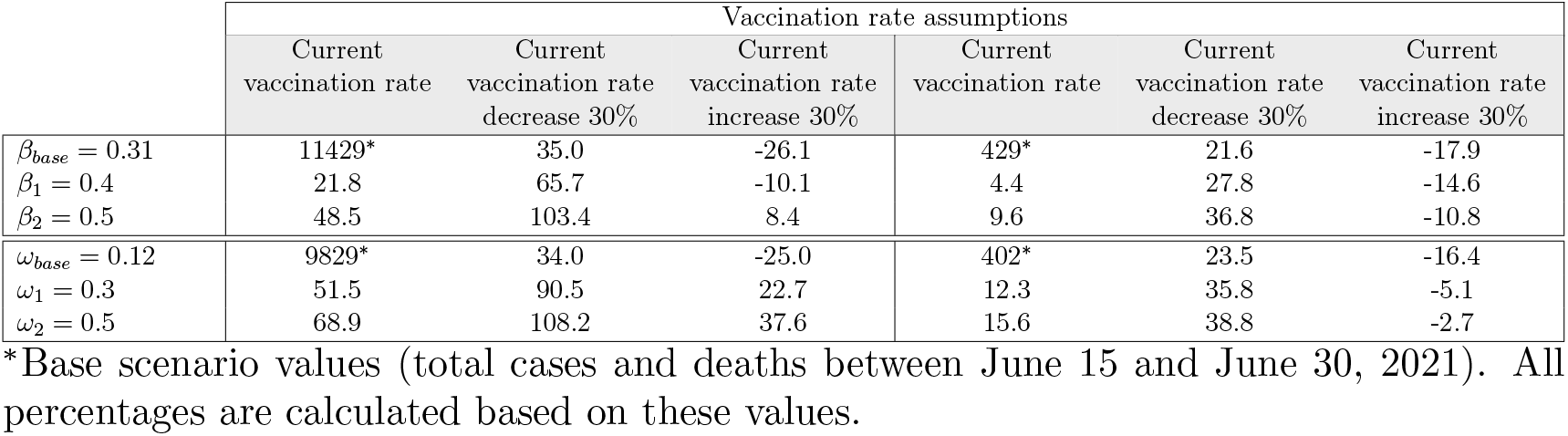
Parameters values for the baseline scenario correspond to the posterior median value *β*_*base*_ = 0.31, *ω*_*base*_ = 0.12, *λ*_*v*1_ = 0.00598, *λ*_*v*2_ = 0.032; *β* = 0.4, 0.5, and *ω* = 0.3, 0.5 were selected according to historical data in CA (Fig S6).

### Vaccine and effective population effects on projections

Projections of confirmed cases and deaths post June 15 that consider scenarios in which the effective population size could increase due to removal of restrictions, resulting in gatherings of susceptible unvaccinated individuals exhibits higher cases and deaths, and much slower downward trends over the period of projection (Fig 2(c), (d)). Increasing the effective population from the baseline of 0.12 to 0.3 resulted in a 51.5% and 12.3% increase in cases and deaths, respectively; these estimates were further increased to 90.5% for cases and 35.8% for deaths with a 30% reduction in the current vaccine uptake.

## Discussion

Our data-driven models produced projections of COVID-19 cases and deaths following the reopening of California’s economy. The real-time Bayesian data assimilation modeling approach provides a more realistic method that leverages historical data. This model takes into account time dependence of contact rates, effective population size, and vaccination rates resulting from the implementation of containment strategies and other factors that have fluctuated throughout this pandemic. We investigated several scenarios to provide insight into the effects of the potential consequences of easing mask-wearing restrictions, decreased social distancing, and the growing threat of coronavirus variants in the changing landscape of vaccine uptake. The baseline scenario in which vaccination rate, transmission rate, and the effective susceptible population stays roughly constant through July, and we found that state-aggregate case and death rates will steadily decrease. However, if removal of restrictions leads to increases in transmission rate or increases in the effective susceptible population, then the current decreasing trend of case rates and death rates is liable to reverse itself, and these rates will persist at a low level. This is especially true if vaccination rates continue to slow from estimates in May. A combination of increase in transmission rates (or effective population) due to relaxing mitigation measures and a decrease in vaccination rates will likely lead to another surge of cases. Our findings suggest that preventive measures with the capacity to impact transmission and the effective susceptible population should be considered, at least until vaccination rates across California reach equitable levels of protection.

Based on data from the current pandemic and information on other related betacoronaviruses, the current dominant strains of SARS-CoV-2 are on track to ultimately become a mild, endemic disease [20]. Unfortunately, that process could take years. A steady but low rate may not initially draw much concern, but the aggregate nature of such data (and derived predictions) can hide some important dynamics. Vaccines have not been equitably dispensed across the state [17], and some counties have consistently had higher case and death rates than others. A stable observed aggregate could very well hide a local outbreak, especially in chronically underserved areas with less robust disease surveillance capabilities. The greater the number of outbreaks and the greater their severity, the more likely a new strain is to emerge.

We assumed that neither natural immunity nor pharmaceutical immunity waned over the analysis window, meaning someone who recovered in April will not have lost any protection by the middle of July. While this fits observations that reinfection is rare in the months after recovery [15], longer-term natural immunity is still unknown. While antibodies for similar betacoronaviruses, such as SARS-CoV-1, become undetectable within a matter of months, T cells remain detectable for a decade or more [21]. There may also be significant cross reactivity with other betacoronaviruses, including the four human coronaviruses that cause the common cold [21] [9], though the SARS viruses seem to create a more specific secondary immune response than the other betacoronaviruses tested [21]. Developing this long-lasting immune response from exposure to a mild disease in childhood can lead to low-impact infections in old age, even if the disease continues to spread [20].

Any exit strategy that aims to push an active infection into an endemic state must account for the possibilities discussed here. There are still situations that lead to consistent low rates of infections, and some that lead to localized or even statewide resurgence. Beyond that, there are key warning signs that this strategy is failing, such as rising reinfection rates, increasing vaccine failure, and rising severity among children. Active monitoring of the situation will be required beyond the date of opening, possibly for years to come.

This work represents the forecast of a dynamic model with eight epidemiological classes and vaccination that aims to simulate the disease transmission process to provide projections of cases and deaths in California after the reopening of its economy. We have demonstrated the potential of a Bayesian data assimilation method to capture the temporal evolution of the parameters involved in the transmission model through a trade-off between the complete history of the outbreak and its latest behavior. This model allows us to capture changes in human behavior, virus dynamics, and restriction measures through the time dependence introduced into the parameters involved in the model each time the prediction is updated. This method uses a data window that moves each time new data is updated using the same number of data for each forecast as well as the number of parameters to estimate. We found that the contact parameters and the effective susceptible population are the most critical parameters that influence the projections of cases and deaths, post reopening.

### Limitations

There are several limitations within our modeling framework that are important to address. Our models do not explicitly capture forms of social influence and individual level behavior which may influence virus spread. Our model assumed homogeneous host mixing which assumes that all participants have identical rates of contacts leading to disease transmission. To mitigate the restrictiveness of this assumption, we included the effective population parameter to our model, which allows for added flexibility in our assumed proportion of individuals susceptible to being infected due to different restrictions, openings, and social behavior. In addition, our model is time dependent allowing the estimated model parameters to vary according to historical data. However, our current model is unable to fully capture the dynamics with specific or localized restrictive measures, or super-spreader events. Another limitation is the absence of age and other risk factors, such as comorbidities, that may impact both infections and hospitalizations. Some of these aspects can be included, as well as more detailed transitions of the dynamics of the virus. However, for practical purposes, our transmission model has made a large number of simplifying assumptions mostly driven by the inability to access data with the appropriate spatio-temporal resolution and coverage. Another limitation in our model is our assumption of the infectivity rate, although currently based on historical data for California, it is likely that it is dynamically changing over time as new variants arise.

Another important limitation in this study pertains to the quality and availability of the data used. The reliability of the projections made by our model depends on the quality of the input data. Our model uses both records of confirmed cases and deaths of COVID-19 due to a belief that the records of deaths are less subject to sampling bias. The case count data depends on the test protocol used in each locality. In most cases, the tests are performed when symptoms appear, introducing a bias due to the growing evidence that asymptomatic individuals are infectious and individuals who eventually become symptomatic and infectious before the onset of any symptoms. In our model, we postulate a value for the proportion of observed and unobserved infectious that depends on the local practices of applying tests and data reporting.

### Data reporting

The databases necessary for the estimation of parameters and code implemented for the study are available in the github repository https://github.com/mdazatorres/Vaccination-Model-for-COVID-19. Analyses were carried out using Python version 3.

## Data Availability

The data underlying the results presented in the study are publicly available and fully anonymized from California's open data portal.

https://data.chhs.ca.gov/dataset/covid-19-time-series-metrics-by-county-and-state

https://data.ca.gov/dataset/covid-19-vaccine-progress-dashboard-data

## Supporting information

### Daily average vaccination rates in California

The average weekly doses of COVID-19 vaccines administered in California have been decreasing since April 11, 2021, coincidentally the week with the highest number of doses administered (Fig S1). In April 11, 2021, the average daily total doses of vaccine administrated were 400,358. The daily average in May 18, 2021, was 222,218. This represents a 44.5% reduction in vaccine uptake compared with the highest rate documented. In the week of April 11, a daily average of 253,785 new doses and 211,898-second doses were administered. In May 18, 2021, the average of first doses administrated decreased by 64% and second doses decreased by 34%, compared to the vaccine rates on April 11. Based on this vaccination trajectory for California, we proposed scenarios on the reduction and uptake of vaccination rates of 30%.

**Figure S1:**
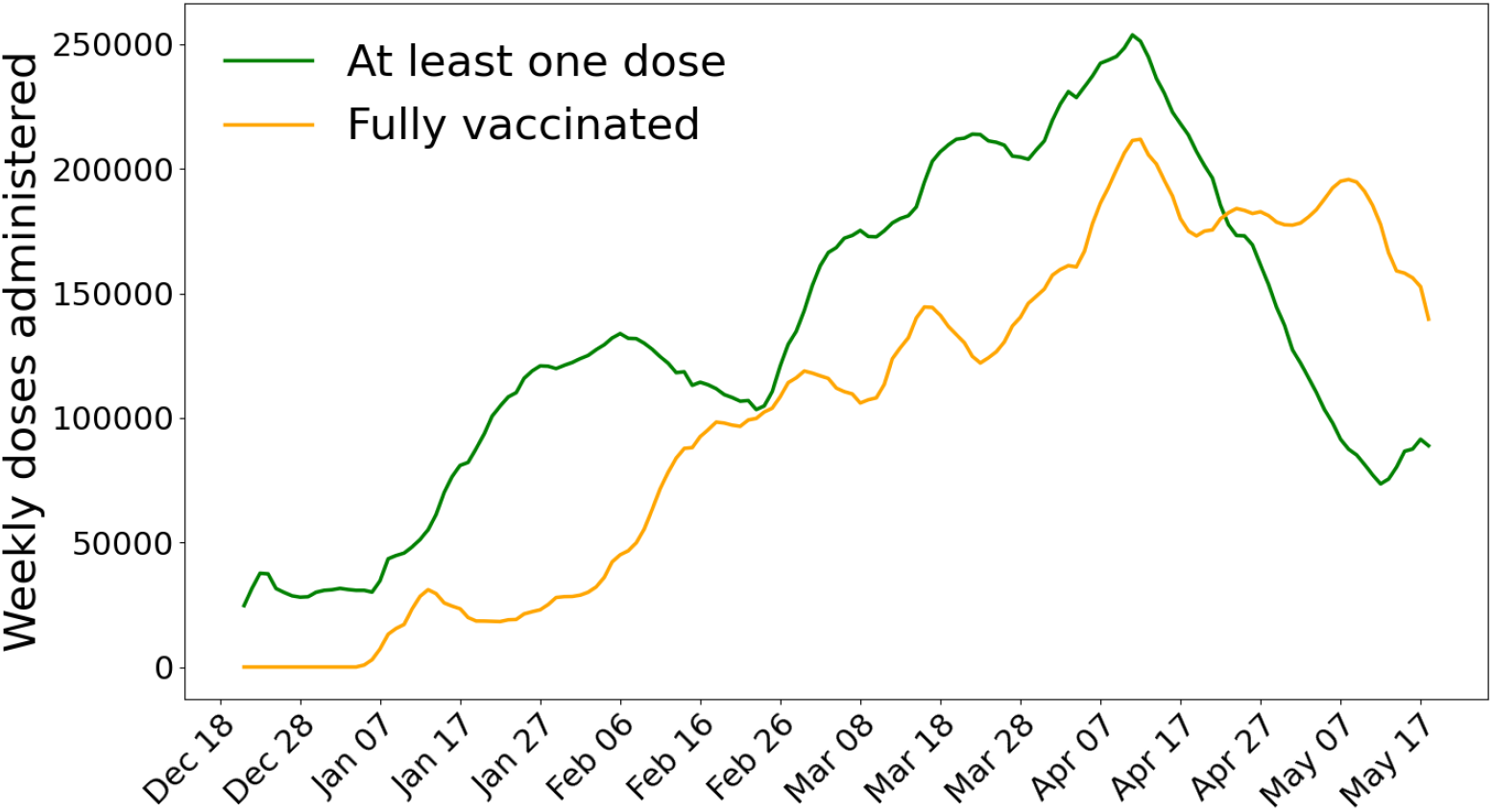
Moving average of administrated doses in California. We use a 7-day moving average to visualize the number of administrated doses. This average is calculated for each day by averaging the values of that day and the six days before. This approach helps prevent significant events (such as changing reporting methods) from skewing the data.

### Bayesian analysis

To conduct parameter estimation, we work with a decoupled model, explained below. Once the parameters are estimated, the model described in the main text (Eq 1) is used to simulate the different scenarios.

### Vaccine coverage model

To model future vaccination coverage, we proposed a compartmental model that includes the dynamics between the not vaccinated population, those who got the first vaccine dose and those who are fully vaccinated. Let *W* be the not vaccinated people at time *t*. We assume that no vaccines have been administered at *t* = 0, which implies *W* (0) = *N*. Then, by assuming that we vaccinate individuals at a constant rate for both first dose and second dose proportional to the current population, we have *W* (*t*), *V*_1_(*t*) and *V*_2_(*t*) that satisfy,

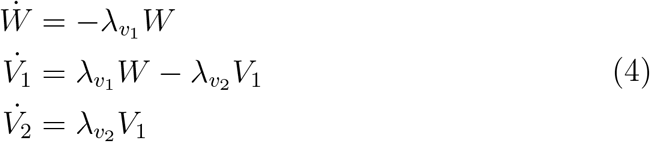

where *W* (0) = *N, V*_1_(0) = 0, and *V*_2_(0) = 0. We stored the cumulative vaccinated population with at least one dose as:

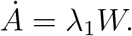

Since real-world vaccination rates 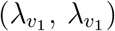 have changed since the start of the vaccination, we used the prior 30 days of real-world vaccination data to adjust our model rates. Information on fully vaccinated people and people with at least one dose is required to find the value of 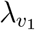 and 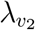.

### Observational model and data

The observed data used to fit the model (4) are based on the records of people with at least one dose and people fully vaccinated. We consider daily data from the first dose of vaccines administered *a*_*i*_ and its theoretical expectation that is estimated in terms of the dynamical model as

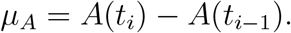

Analogously, we consider daily second doses administered *u*_*i*_, and its theoretical expectation given by

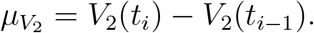

### Estimation of model parameters with MCMC

To carry out a likelihood-based analysis, we postulate that the number of both at least one dose and fully vaccinated doses administered follows a Poisson distribution, *Pois*. For data, *y*_*i*_, we let

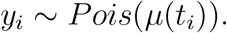

We assume conditional independence in the data, therefore from the Poisson model, we obtain a likelihood. Our parameters are 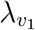 and 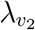. Regarding the elicitation of the parameters prior distribution, we use a Gamma distribution with scale 3 and shape parameter 10. To sample from the posterior, we resort to MCMC using “t-walk” generic sampler [11].

### Posterior distribution and vaccine coverage model results

Estimation of vaccination rates is used to predict vaccine coverage as well as the dynamics of SARS-CoV-2. We displayed the results using the data reported 30 days prior of May 18, 2021. We ran 10,000 iterations of the t-walk and after a burn-in period of 1,000 iterations, the chain seems to be sampling from the equilibrium distribution (i.e., the posterior distribution) (Fig S2(c)-(d)). Fig S2(a)-(b) correspond to the marginal posterior distribution for 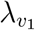 and 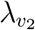. The histograms are reported with 9,000 samples since the first (burn-in) 1,000 are discarded.

**Figure S2:**
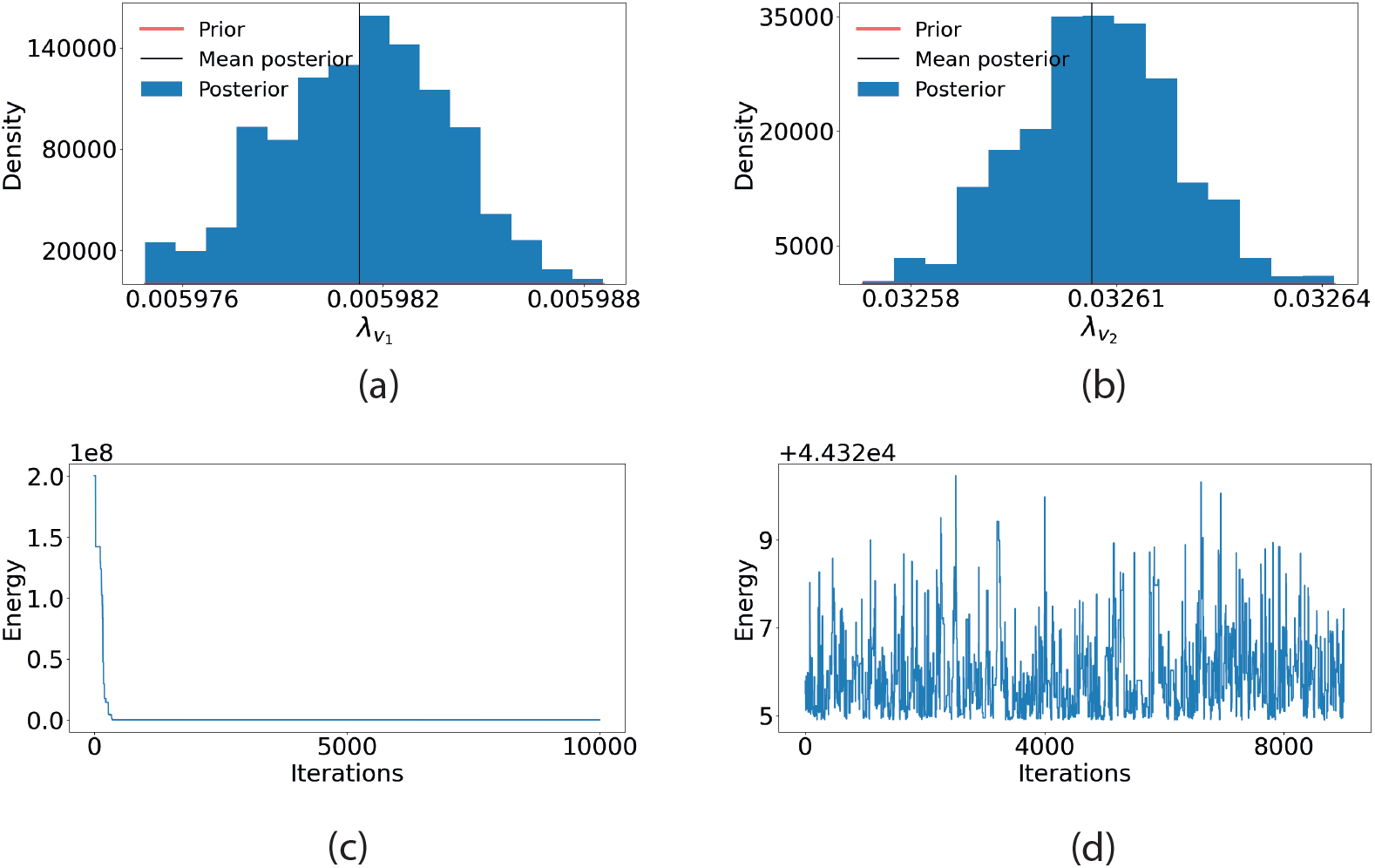
Marginal posterior. After 10,000 MCMC samples, (a) the marginal posterior distribution for the vaccinate rates (b) 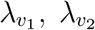 and its prior distribution (red). (c) Trace plot of the logarithm of the posterior distribution and (d) Trace plot of the logarithm of the posterior distribution without a burn-in of 1,000 iterations.

**Figure S3:**
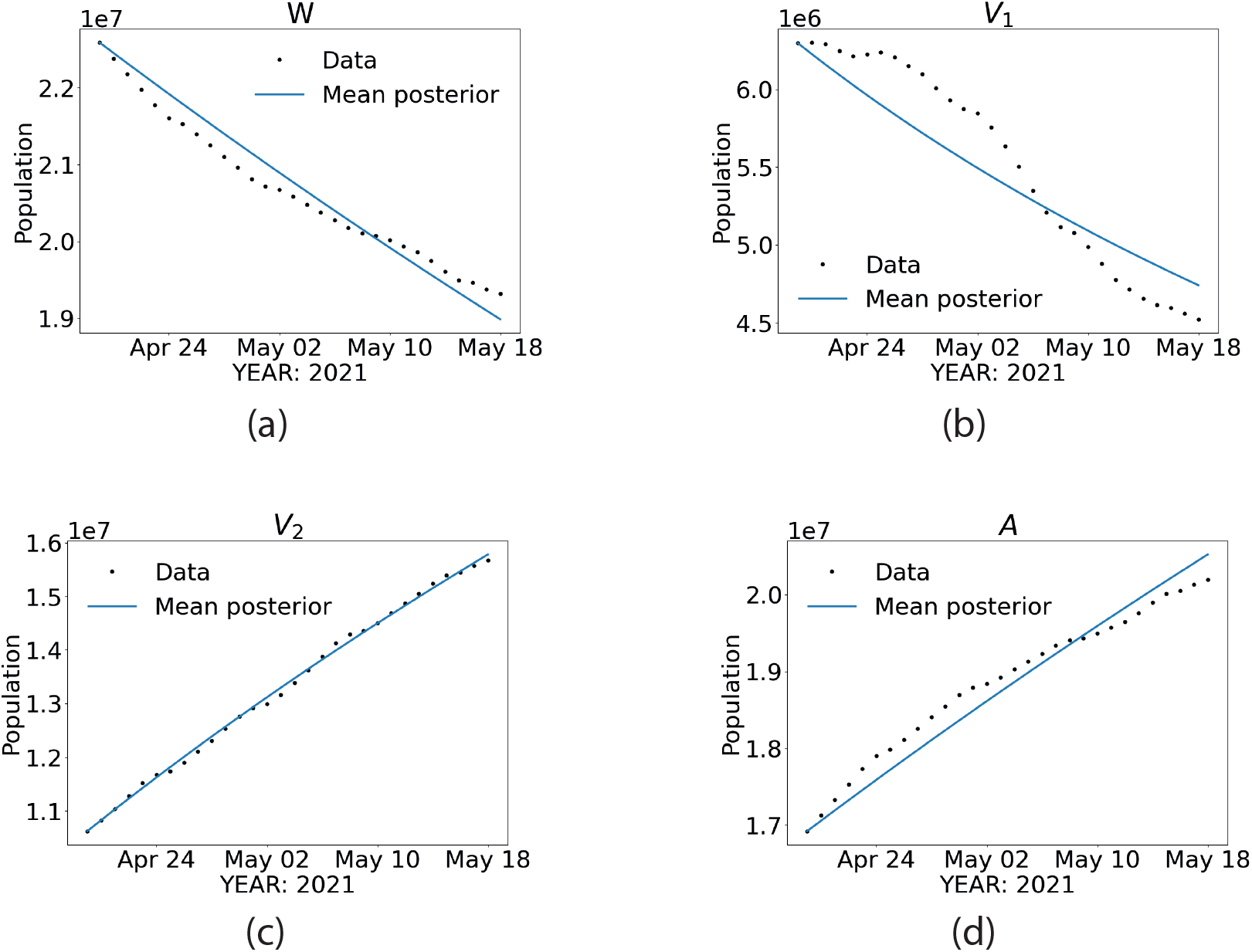
Vaccine coverage model trajectory with median posterior values. (a) people not vaccinated, (b) people with at least one dose, (c) fully vaccinated, and (d) cumulative total of people taking at least one dose at time. *t*.

### Transmission model

To have more realistic scenarios, we estimate the parameters involved in the dynamical of the SARS-CoV-2 transmission using the Bayesian Sequential Forecasting Method (BSFM) proposed in [13].

### Bayesian Sequential Forecasting Method

The BSFM updates the evolution of the dynamic system from the posterior distribution of both model parameters and state variables as new epidemic records become available. New prior models are defined from the current parameters and state variables posterior distributions on a sliding time window.

Within each sliding window, posterior distributions are computed beyond the available epidemic records to produce the forecasts.

Let *x*(*t*) = (*S*(*t*), *E*(*t*), *I*(*t*), …)^*T*^ denote the time–dependent vector of state variables. We shall assume that the epidemic and transmission models are coded in a dynamic system

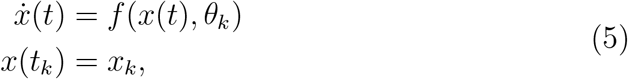

where *t*_*k*_ and *x*_*k*_ denote the initial time and state in the forecasting window [*t*_*k*_, *t*_*k*_ + *L* + *D* + *F*] respectively, *θ*_*k*_ is a vector of model parameters (e.g., contact rate *β*, effective population size *ω*, etc.) used to calibrate the model (5), *L* is the learning period size, *F* is the number of days to forecast, *n* is the number of days to move the forecasting window, and *D* is the number of the delays days. We denoted *p*^(*k*)^ := (*x*_*k*_, *θ*_*k*_) as the joint vector of initial conditions and model parameters to be inferred.

The initial forecasting, *k* = 0, is done as a usual Bayesian inference problem, we postulate

- A prior distribution, 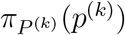.
- A likelihood, 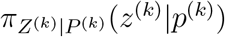, where *z*_*k*_ represents epidemic records in *t*_*k*_ to *t*_*k*+1_ + *L* (e.g., confirmed cases, deaths, etc.).
- We use equation (5) and samples obtained through Markov Chain Monte Carlo of the corresponding posterior distribution, *π*_*P* |*Z*_(*p*^(*k*)^ |*z*^(*k*)^) to make a probabilistic prediction of *x*(*t*) in the forecasting period *t* ∈ [*t*_*k*_ + *L* + *D, t*_*k*_ + *L* + *D* + *F*].

To the next forecasting *k >* 0, we update the forecasting window by setting *t*_*k*+1_ = *t*_*k*_ + *n*. The new forecasting window is [*t*_*k*+1_, *t*_*k*+1_ + *L* + *D* + *F*]. The prior distribution of *p*^(*k*)^ = (*x*_*k*_, *θ*_*k*_) is set using the MCMC output of the period *k* − 1:

- For the *k*−initial state (*x*_*k*_), the MCMC output of the state variable *x*(*t*) at time *t*_0_ + *nk* obtained with equation (5) is fitted with a known distribution.

**Figure S4:**
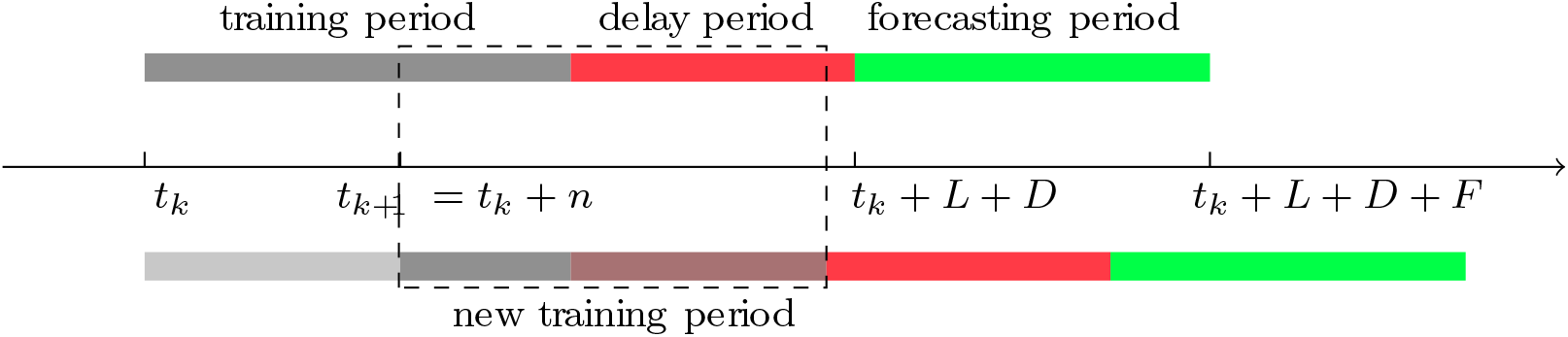
Bayesian Sequential data assimilation.
- For the model parameters *θ*_*k*_, the MCMC output of *θ*_*k−*1_ is fitted a known distribution.
- Finally, we set *k* ←*k* + 1 and repeat the above process to create a new forecast.

The central part of BSFM is that the time dependence of the transmission model parameters is introduced by updating sequential forecasts reported on the history of the outbreak using the posterior distributions as prior distributions for the parameters in the current forecast. Thus, our transmission model becomes a non-autonomous dynamic system, with the same amount of parameters in time and data, capable of capturing changes in outbreak behavior produced by human and virus trend changes.

The Bayesian filtering method predicts along the dynamical system (5) evaluated in sample points of the posterior distribution 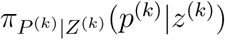 in the current forecasting window [*t*_*k*_, *t*_*k*_ + *L* + *D* + *F*].

### Application to California Data

The parameter estimation is carried out with model [6], without the vaccination transitions.

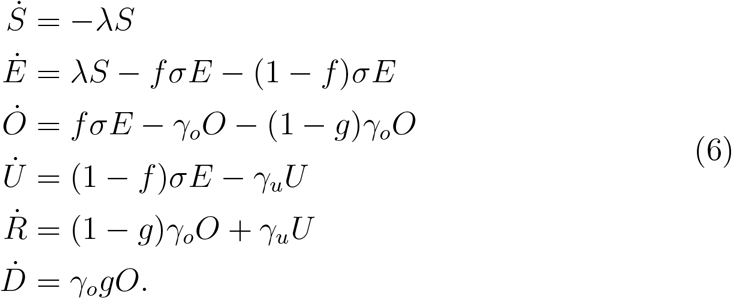

Using data of confirmed cases and deaths, we estimate the contact rate (*β*), the proportion of the effective population (*ω*), the fraction of individuals infected that are deceased (*g*), and the initial conditions for all compartments, except for the susceptible ones, which are set as *S*(*t*_0_) = *ω* · *N* − (*E*(*t*_0_) + *O*(*t*_0_) + *U*_(_*t*_0_) + *R*_(_*t*_0_)) + *V*_1_(*t*_0_) + *V*_2_(*t*_0_).

We consider records of confirmed cases and deaths from January 25, 2020, until May 10, 2021. These data are smoothing using a weekly moving average, which is calculated for each day by averaging the values of that day and the six days before. This approach helps prevent significant events (such as changing reporting methods) from skewing the data.

Using the BFSM for California data, we forecast every eight days with the most recent data. Fig S5 shows the forecasts from December 6 to May 18, 2021, for both confirmed cases and deaths. Fig S6 shows the trajectory of *ω, θ*, and *g* for the pandemic period. The public response to long-term mitigation measures for the pandemic is reflected in the evolution of *β* and *ω* parameters.

The *β* contact rate takes a high value at the beginning of the pandemic but declines after the first intervention in California (March 12, 2021) and stabilizes around 0.35. The value of *ω* has shown greater variability over time. Like *β*, it takes high values at the beginning of the pandemic and declines with the first intervention carried out in California in March. In July and December, we observed an increase in the values of this parameter that coincides with the waves that California had in the same months. The proportion of observed deaths (*g*) decreases with time, probably due to experience gained over time in caring for patients and expanding hospital capacity. After a while, this value stabilizes and remains close to 0.03.

For the vaccination model (Eq 4 main text), we use the posterior median in the last forecast for initial conditions, contact rate *β*, effective population *ω*, and proportion of deaths *g*. We take the initial condition for vaccination like the current vaccine and redefine *S*(*t*_0_) = *ωN*_0_−*V*_0_−*E*_0_−*U*_0_−*O*_0_−*R*_0_−*D*_0_.

**Figure S5:**
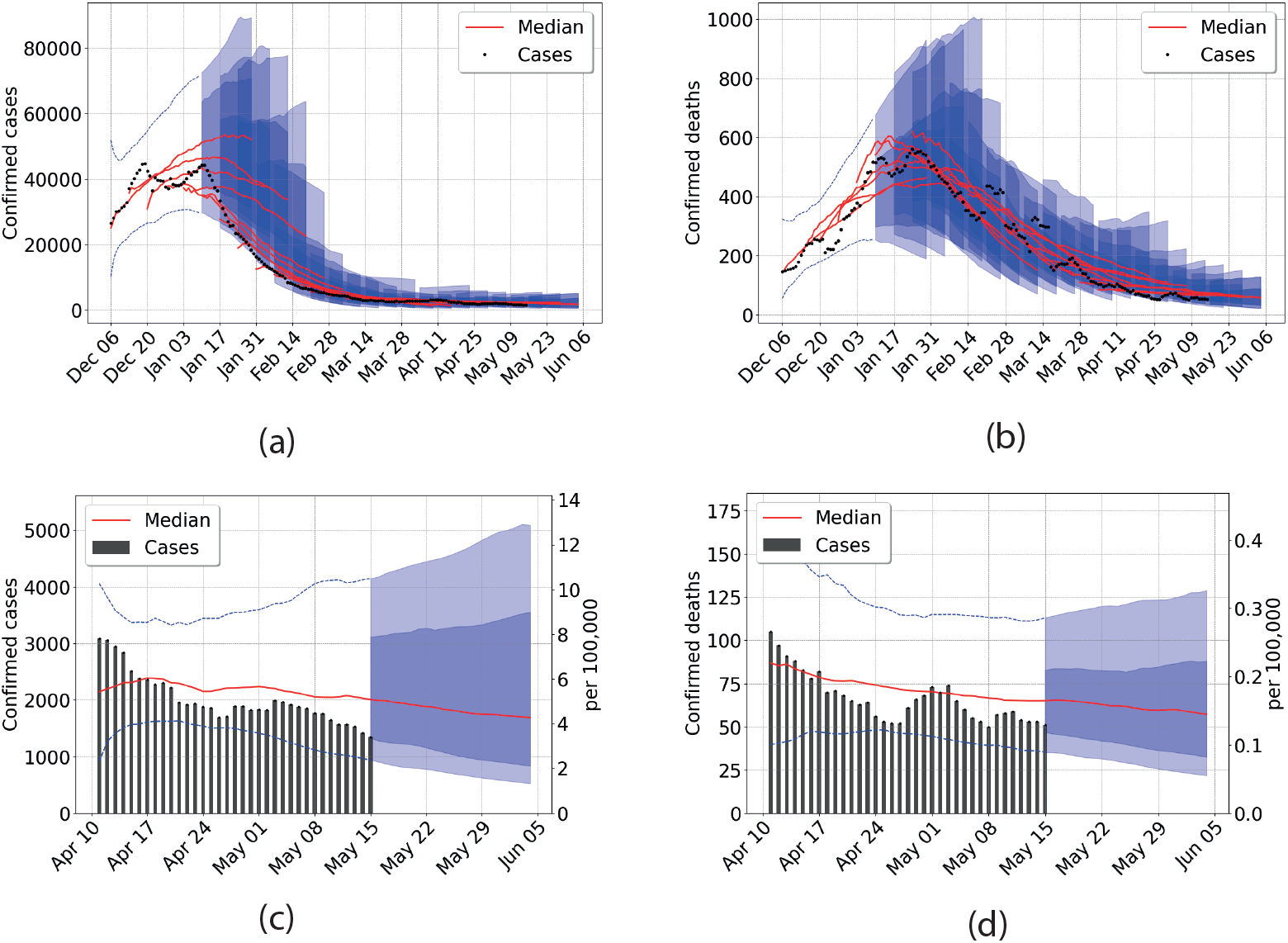
California outbreak analysis. Data from April 8 to May 18, 2021 is used. (a) Confirmed cases (b) Confirmed deaths. Central red lines indicate the median incidence forecast. The darker shaded region indicates the interquartile forecast range, and the lighter shaded region indicates the 5–95th percentile range. All displayed forecast duration’s are ten days from the point of prediction. California total population 39,512,223.

### Prediction interval coverage

We have the probabilistic one-, two-, three-, and four-week ahead forecasts of the total number of confirmed cases and deaths due to COVID-19 from January 13, 2020, to May 18, 2021, and every eight days after that for California. We evaluated our forecast error with the calibration of the prediction interval coverage (80% and 50%). The prediction interval coverage is calculated by determining the frequency with which the prediction interval contained the eventually observed outcome. We do this for all prediction intervals calculated from January 13, 2020, to May 18, 2021, and calculate the average of these. In a model that accurately characterizes uncertainty, the prediction interval level will correspond closely to the frequency of eventually observed outcomes that fall within that prediction interval. For example, finally observed values should be within the 50% prediction interval approximately 50% of the time.

**Figure S6:**
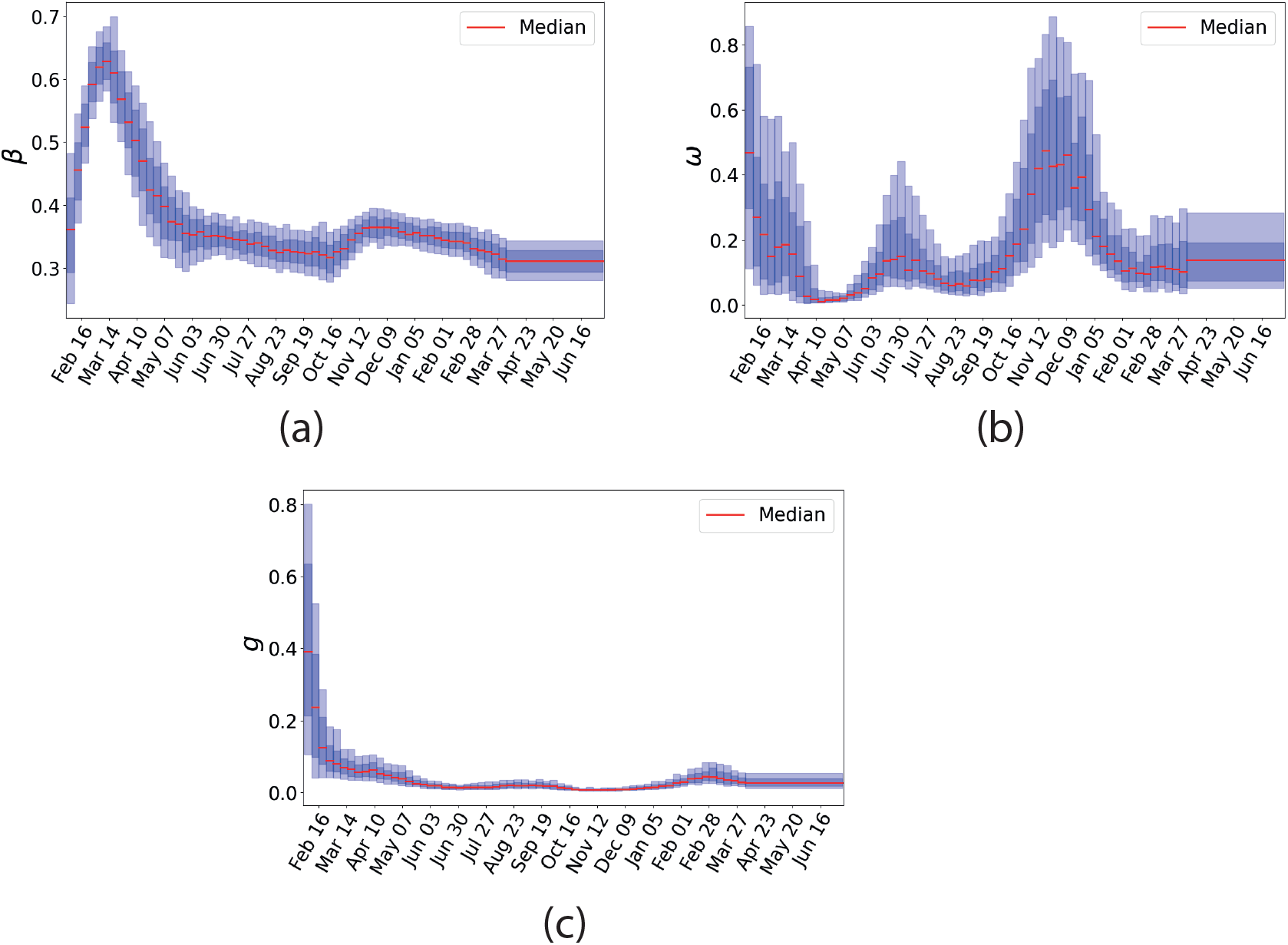
The trajectory of the parameters during the pandemic period. From left to right, contact rate after lockdown (*β*), proportion of the effective population (*ω*), and the fraction of infected dying (*g*). Central red lines indicate median incidence forecast. Darker shaded region indicates forecast interquartile range, and lighter shaded region indicates 5–95th percentile range.

**Table S1:** Observed prediction interval coverage.

| Measure | Cases |  |  |  | Deaths |  |  |  |
| --- | --- | --- | --- | --- | --- | --- | --- | --- |
|  | Observed Prediction Interval<br>Forecast Horizon (weeks ahead) |  |  |  | Observed Prediction Interval<br>Forecast Horizon (weeks ahead) |  |  |  |
|  | week 1 | week 2 | week 3 | week 4 | week 1 | week 2 | week 3 | week 4 |
| 50% Coverage | 0.60 | 0.56 | 0.52 | 0.49 | 0.59 | 0.64 | 0.66 | 0.67 |
| 80% Coverage | 0.83 | 0.80 | 0.76 | 0.73 | 0.94 | 0.94 | 0.93 | 0.91 |

The forecasts were well-calibrated, with prediction intervals covering the observed data with the expected frequency (Table S1). The 50% prediction intervals captured 48-60% of observations for all forecast horizons for confirmed cases and 58-66% for deaths. The 80% prediction intervals captured only 73-83% of confirmed cases and 91-94% for deaths. The intervals were better calibrated for confirmed death due that the record of Covid-19 deaths is more reliable than records of confirmed cases. The last one depends directly on the number of tests applied.

## Other Results

These results are similar to the results described in the main text, but instead of reducing the current vaccination rate by 30%, we consider the scenario where this value decreases by 60%.

**Figure S7:**
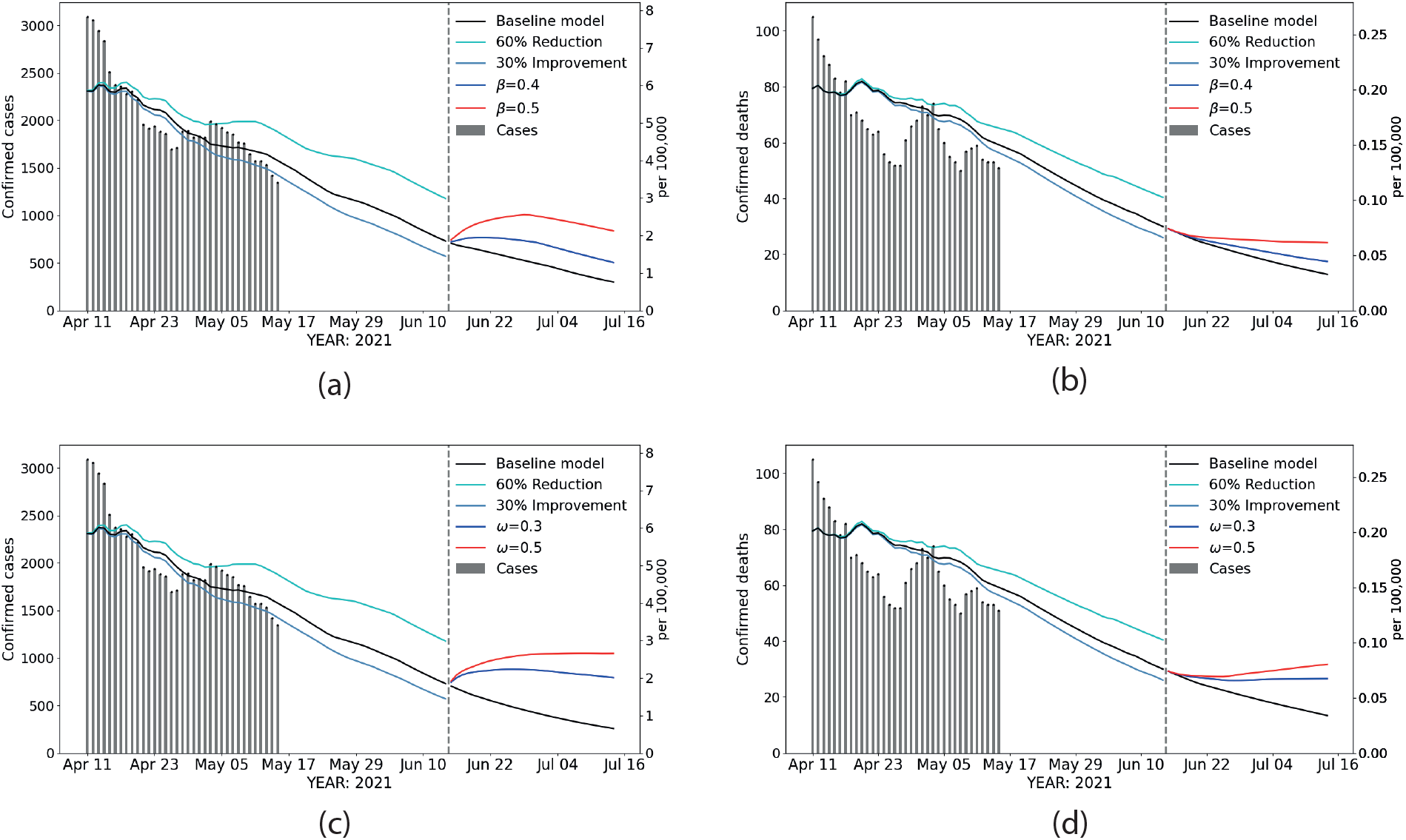
Scenarios. The dashed vertical line indicates the opening day, June 15. Reported data are shown in gray (bars). The black line corresponds to the baseline scenario, the cyan line corresponds to the projection assuming a 60 % decrease in the current vaccination rate, and the magenta line corresponds to the projection assuming a 30% increase in the current vaccination rate. After June 15, the red, blue, and black lines correspond to the projection with the different values of *ω*, cases (a) and deaths (b) and changes in *β* (c) cases and (d) deaths, with the current vaccination rate.

**Table S2:** Parameters values for the baseline scenario correspond to the posterior median value *β*_*base*_ = 0.31, *ω*_*base*_ = 0.12, *λ*_*v*1_ = 0.00598, *λ*_*v*2_ = 0.032; *β* = 0.4, 0.5, and *ω* = 0.3, 0.5 were selected according to historical data in CA (Fig. S6).

| Parameters | Vaccination rate assumptions |  |  |  |  |  |
| --- | --- | --- | --- | --- | --- | --- |
|  | Current vaccination rate | Current vaccination rate decrease 60% | Current vaccination rate increase 30% | Current vaccination rate | Current vaccination rate decrease 60% | Current vaccination rate increase 30% |
|  | Increase or prevention percentage in cases 15 days after opening |  |  | Increase or prevention percentage in deaths 15 days after opening |  |  |
| $\beta_{base} = 0.31$ | 11429* | 80.4 | -26.1 | 429* | 50.7 | -17.9 |
| $\beta_1 = 0.4$ | 21.8 | 122.6 | -10.5 | 4.4 | 61.1 | -14.6 |
| $\beta_2 = 0.5$ | 48.5 | 172.5 | 8.4 | 9.6 | 73.5 | -10.8 |
| $\omega_{base} = 0.12$ | 9829* | 77.5 | -25.6 | 402* | 52.0 | -16.4 |
| $\omega_1 = 0.3$ | 51.5 | 139.1 | 22.7 | 12.2 | 65.8 | -5.1 |
| $\omega_2 = 0.5$ | 68.9 | 159.9 | 37.6 | 15.6 | 69.2 | -2.7 |

\*Base scenario values (total cases and deaths between June 15 and June 30, 2021). All percentages are calculated based on these values.

## Notes

### Competing Interest Statement

The authors have declared no competing interest.

### Funding Statement

There is no funding to report.

## References

[1] 10 reasons why pandemic fatigue could threaten global health in 2021 — gavi, the vaccine alliance. https://www.gavi.org/vaccineswork/10-reasons-why-pandemic-fatigue-could-threaten-global-health-2021. (accessed on 05/27/2021).

[2] California Department of Public Health. Beyond the Blueprint For a Safer Economy.

[3] California Department of Public Health. Blueprint For a Safer Economy. https://covid19.ca.gov/safer-economy/. (accessed 5/20/21).

[4] California Open Data Portal. COVID-19 Vaccine Progress Dashboard Data. https://data.ca.gov/dataset/covid-19-vaccine-progress-dashboard-data. (accessed on 05/17/2021).

[5] Covid-19 variants and vaccines: 9 questions about coronavirus variants, answered - vox. https://www.vox.com/22385588/covid-19-vaccine-variant-mutation-n440k-india-moderna-pfizer-b1617. (accessed on 05/27/2021).

[6] New variants of coronavirus: What you should know — johns hopkins medicine. https://www.hopkinsmedicine.org/health/conditions-and-diseases/coronavirus/a-new-strain-of-coronavirus-what-you-should-know. (accessed on 05/27/2021).

[7] U.S. Census Bureau. Quick Facts: California. https://www.census.gov/quickfacts/CA. (accessed on 05/19/2021).

[8] What the science says about lifting mask mandates. https://www.nature.com/articles/d41586-021-01394-0. (accessed on 05/27/2021).

[9] Dvir Aran, Daniel C. Beachler, Stephan Lanes, and J. Marc Overhage. Prior presumed coronavirus infection reduces covid-19 risk: A cohort study. Journal of Infection, 81:922–930.

[10] Qifang Bi, Yongsheng Wu, Shujiang Mei, Chenfei Ye, Xuan Zou, Zhen Zhang, Xiaojian Liu, Lan Wei, Shaun A Truelove, Tong Zhang, et al. Epidemiology and transmission of covid-19 in shenzhen china: Analysis of 391 cases and 1,286 of their close contacts. MedRxiv, 2020.

[11] J Andrés Christen, Colin Fox, et al. A general purpose sampling algorithm for continuous distributions (the t-walk). Bayesian Analysis, 5(2):263–281, 2010.

[12] Noa Dagan, Noam Barda, Eldad Kepten, Oren Miron, Shay Perchik, Mark A Katz, Miguel A Hernán, Marc Lipsitch, Ben Reis, and Ran D Balicer. Bnt162b2 mrna covid-19 vaccine in a nationwide mass vaccination setting. New England Journal of Medicine, 384(15):1412–1423, 2021.

[13] Maria L Daza-Torres, Marcos A Capistrán, Antonio Capella, and J Andrés Christen. Bayesian sequential data assimilation for covid-19 forecasting. arXiv preprint 2103.06152, 2021.

[14] Centers for Disease Control and Prevention. Covid-19 pandemic planning scenarios — cdc. https://www.cdc.gov/coronavirus/2019-ncov/hcp/planning-scenarios.html#table-2. (accessed on 03/11/2021).

[15] Aidan T. Hanrath, Brendan A.I. Payne, and Christopher J.A. Duncan. Prior sars-cov-2 infection is associated with protection against symptomatic reinfection. Journal of Infection, 82:E29–E30.

[16] California Health and Human Services Open Data Portal. Covid-19 time-series metrics by county and state - datasets. https://data.chhs.ca.gov/dataset/covid-19-time-series-metrics-by-county-and-state. (accessed on 06/01/2021).

[17] MM Hughes, A Wang, MK Grossman, and et al. County-Level COVID-19 Vaccination Coverage and Social Vulnerability — United States, december 14, 2020–march 1, 2021. MMWR Morb Mortal Wkly Rep, 70:431–436, 2021.

[18] Jocelyn Keehner, Lucy E. Horton, Michael A. Pfeffer, Christopher A. Longhurst, Robert T. Schooley, Judith S. Currier, Shira R. Abeles, and Francesca J. Torriani. Sars-cov-2 infection after vaccination in health care workers in california. New England Journal of Medicine, 384(18):1774–1775, 2021.

[19] Kai Kupferschmidt. The pandemic virus is slowly mutating. but does it matter?, 2020.

[20] Jennie S. Lavine, Ottar N. Bjornstad, and Rustom Antia. Immunological characteristics govern the transition of covid-19 to endemicity. Science, 371(6530):741–745, 2021.

[21] Nina Le Bert, Anthony T. Tan, Kamini Kunasegaran, Christine Y. L. Tham, Morteza Hafezi, Adeline Chia, Melissa Hui Yen Chng, Meiyin Lin, Nicole Tan, Martin Linster, Wan Ni Chia, Mark I-Cheng Chen, Lin-Fa Wang, Eng Eong Ooi, Shirin Kalimuddin, Paul Anantharajah Tambyah, Jenny Guek-Hong Low, Yee-Joo Tan, and Antonio Bertoletti. Sars-cov-2-specific t cell immunity in cases of covid-19 and sars, and uninfected controls. Science, 584:457–462, 2020.

[22] Quan-Xin Long, Xiao-Jun Tang, Qiu-Lin Shi, Qin Li, Hai-Jun Deng, Jun Yuan, Jie-Li Hu, Wei Xu, Yong Zhang, Fa-Jin Lv, et al. Clinical and immunological assessment of asymptomatic sars-cov-2 infections. Nature medicine, 26(8):1200–1204, 2020.

[23] CDC COVID-19 Vaccine Breakthrough Case Investigations Team. Covid-19 vaccine breakthrough infections reported to cdc—-united states, january 1–april 30, 2021. MMWR Morb Mortal Wkly Rep, 2021.

[24] Robert Verity, Lucy C Okell, Ilaria Dorigatti, Peter Winskill, Charles Whittaker, Natsuko Imai, Gina Cuomo-Dannenburg, Hayley Thompson, Patrick GT Walker, Han Fu, et al. Estimates of the severity of coronavirus disease 2019: a model-based analysis. The Lancet infectious diseases, 2020.

